# Maternal complications after cesarean section by obstetric facility type in Japan: A claims-based cohort study

**DOI:** 10.64898/2026.08.12.26360328

**Authors:** Takashi Yoshimasu, Kazuhiro Abe, Misuzu Sato, Kazuki Ohashi, Tasuku Inao, Sachiko Ono, Isao Yokota, Katsuhiko Ogasawara

**Affiliations:** Department of Biostatistics, Hokkaido University; Department of Epidemiology, University of Pittsburgh; Department of Health Care Policy, Hokkaido University; Division of Dental Public Health, Hokkaido University; Faculty of Health Sciences, Hokkaido University; Department of Molecular Psychoneuroimmunology, Hokkaido University; Department of Eat-loss Medicine, Graduate School of Medicine, The University of Tokyo; Faculty of Engineering, Muroran Institute of Technology

**Keywords:** cesarean section, insurance claims data, maternal complication, obstetric facility, population-based study

## Abstract

**Aim:** Little is known about patient safety in a less consolidated obstetric system where various facilities, such as perinatal medical centers (PMCs), general hospitals, and clinics, collaborate under risk-based role differentiation. We aimed to compare maternal complications after cesarean section by facility type across area types (rural, provincial, and metropolitan) in Hokkaido, Japan.

**Methods:** This retrospective cohort study used insurance claims data from Hokkaido (2018–2025). Two comparisons were conducted for a composite outcome of postpartum hemorrhage, infection, and thrombosis: a two-category comparison (PMC vs. non-PMC, combining general hospitals and clinics) across all areas, with an interaction term between facility and area type; a three-category comparison (PMC vs. general hospital vs. clinic) restricted to metropolitan areas. Generalized estimating equations with a Poisson distribution, accounting for clustering within facilities, were applied to estimate risk ratios.

**Results:** A total of 1,822 participants underwent cesarean section. PMCs were associated with lower maternal complication rates compared to non-PMC facilities (adjusted RR 0.37, 95% CI 0.16–0.88 in rural areas; adjusted RR 0.20, 95% CI 0.11–0.37 in provincial areas). In metropolitan areas, PMCs and general hospitals were associated with lower maternal complication rates compared to clinics (PMC vs clinic: adjusted RR 0.42, 95% CI 0.18–0.97; general hospital vs clinic: adjusted RR 0.25, 95% CI 0.09–0.70).

**Conclusions:** Higher-level facilities were associated with lower maternal complication rates after cesarean section in Japan. These findings provide important evidence for regional consolidation of obstetric care.

## Introduction

The number of obstetric facilities has been declining in many countries, resulting in fewer yet larger facilities [1–5]. While consolidation of obstetric care may impair healthcare access, the creation of higher-level, higher-volume facilities may contribute to patient safety, particularly for high-risk procedures such as cesarean section (CS). For instance, higher-volume facilities are associated with lower surgical complication rates, potentially reflecting greater human resources, greater subspecialty availability, and more consistent postoperative care [6]. Similarly, in obstetric care, higher-volume facilities (i.e., greater annual births) are associated with fewer maternal complications [7–9], suggesting that consolidated perinatal healthcare systems may benefit the safety of pregnant individuals.

In Japan, however, obstetric care has long maintained a less consolidated system, where various facilities, including perinatal medical centers (PMCs), general hospitals, and clinics, collaborate under risk-based role differentiation [10]. In particular, PMCs are designated core facilities that receive emergency referrals and high-risk pregnancies from general hospitals and clinics [11]. The strong collaboration between these facilities has long contributed to the low maternal and infant mortality rates [10]. Yet the number of obstetric facilities in Japan has decreased from 3,000 in 2005 to 2,000 in 2024, reflecting the declining birth rates and maldistribution of obstetricians [10]. To sustain healthcare systems amid this ongoing decline in obstetric facilities, Japan’s Ministry of Health, Labour and Welfare (MHLW) has been promoting the consolidation of obstetric care, with PMCs positioned as core facilities in each region [12]. The MHLW has also emphasized regional variations in geography, size, transportation, population, and obstetrician distribution [12]. Consolidation strategies should therefore be tailored to each region’s circumstances.

However, research on regional consolidation is limited, particularly with respect to patient safety within a less consolidated obstetric care system. On average, each facility has only 400–500 annual births in Japan [13], figures substantially smaller than those seen at facilities in other countries, such as the United States, the United Kingdom, Sweden, Portugal, and Finland [2,5,14]. Previous studies from these countries have compared maternal and infant outcomes between low- and high-volume facilities [1,8,9], and few have examined outcomes within low-volume facilities (e.g., annual births <1000). A recent study showed that higher hospital acuity, defined as higher proportions of high-risk pregnancies, was associated with lower maternal end-organ damage after adjusting for the number of deliveries per hospital [15]. This suggests that higher-level facilities may have lower complication rates even among low-volume facilities. Nonetheless, regional variations have rarely been the focus of these studies. Previous studies found that higher facility volume was associated with lower rates of postpartum hemorrhage (PPH) in rural areas, but sometimes higher rates of PPH in urban areas [16,17], highlighting the need to incorporate regional variations into safety assessment. Taken together, evidence remains critically insufficient among low-volume facilities, particularly when accounting for regional variations.

Hokkaido is the northernmost prefecture of Japan, with a population of approximately 5 million and 20,000–30,000 annual births [18]. It encompasses diverse regional variations, including rural, provincial, and metropolitan areas [19]. We particularly focus on CS as it carries a high risk of maternal complications [20] and accounts for 42% of maternal deaths despite a CS rate of approximately 20% in Japan [12,21]. In Japan’s risk-based referral system, facility type (i.e., PMC, general hospital, and clinic) is considered to be more closely linked to patient outcomes than facility volume. Therefore, we aimed to compare maternal complications after CS among facility types across different regions in Hokkaido, Japan.

## Methods

### Study design, setting, and participants

This retrospective cohort study was conducted using medical insurance claims data (April 2018 to October 2025) from all municipalities in Hokkaido. In Japan, all residents are registered in the health insurance system, including national health insurance (NHI, for the self-employed), employee’s health insurance, and a special scheme for the aged (75 years old and older) [22]. NHI data were used for the analyses. NHI covers approximately 25% of the population nationwide [23]. The database includes demographic data, clinical examination, pharmacy claims, medical procedure and device claims, diagnosis codes, and facility codes for each procedure, but does not include laboratory data, the amount of bleeding, coagulation status, or imaging findings. We used MHLW codes because these codes are standardized across different facility payment systems.

The design diagram is shown in Figure 1. The study entry date was defined as the date of CS. The covariate assessment window was defined as the 3-month period preceding the month of study entry, excluding the month of study entry itself. The follow-up window was defined as the 30 days following the study entry date. The exclusion assessment window was defined as the whole period including the covariate assessment and follow-up window.

**Figure 1.**
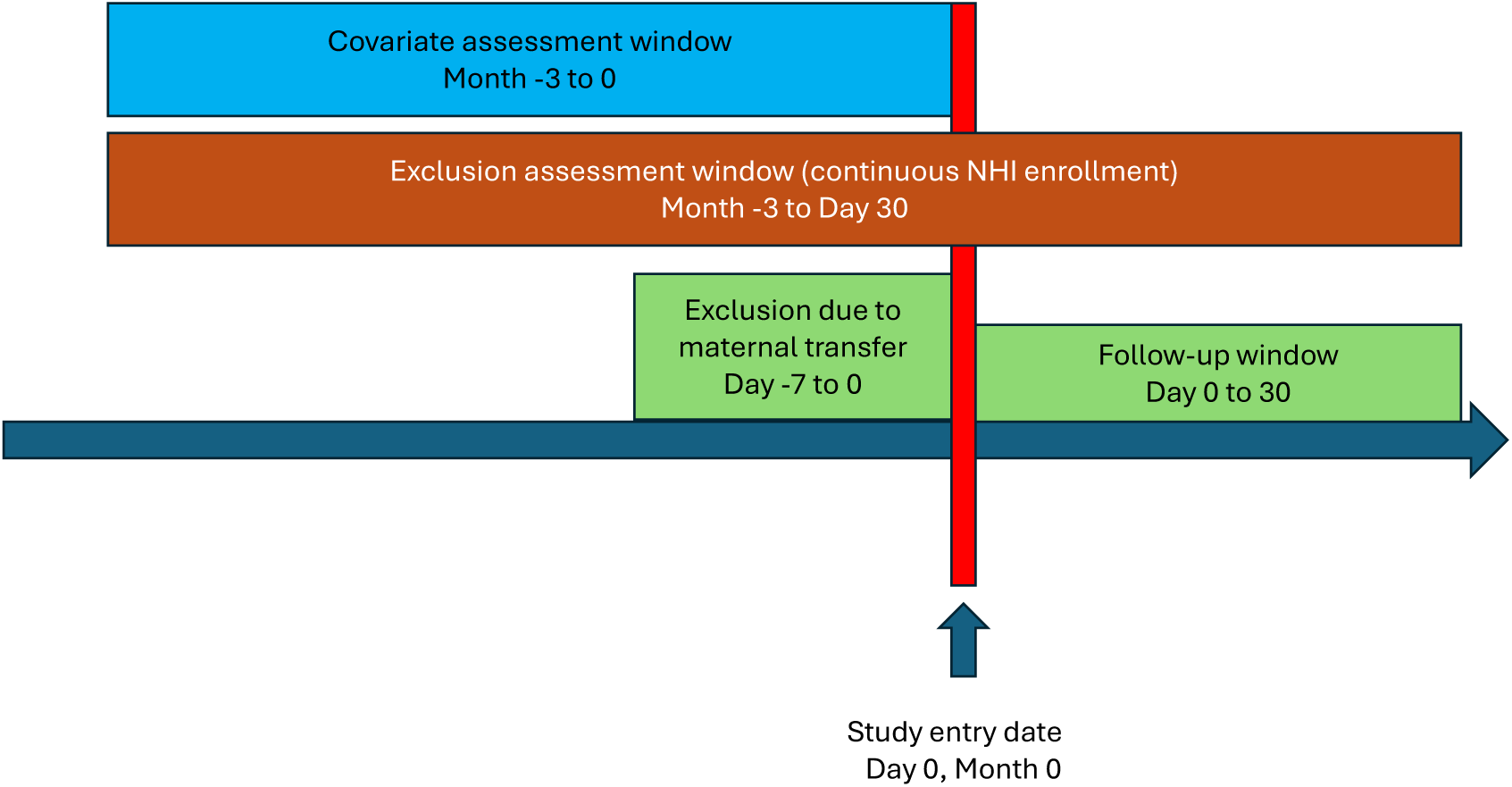
The design diagram.

The study population was pregnant individuals who underwent CS. Exclusion criteria were as follows: (1) lack of continuous enrollment in NHI throughout the exclusion assessment window, (2) maternal transfer within 7 days prior to the study entry date, or (3) serious maternal comorbidity in the components of maternal comorbidity index (described below), such as multiple gestation and placenta previa. These conditions are summarized in **Appendix Table 1.**

The study was approved by the Hokkaido University ethics review committee (Department of Health Sciences, No. 25-62-1). All of the MHLW codes are provided in **Appendix Tables 2–6.**

### Variables

#### Facility type

A hospital is defined as a medical facility with ≥20 beds, and a clinic as one with <20 beds. Among hospitals, PMCs are designated by the government and were identified using facility codes. There are currently 36 PMCs in Hokkaido [24]. First, we examined a two-category comparison (PMC vs. non-PMC, combining general hospitals and clinics) in all area types. Second, we used a three-category comparison (PMC vs. general hospital vs. clinic) restricted to metropolitan areas.

#### Outcome

The outcome was maternal complications, defined as a composite of PPH, infection, and thrombosis [7,25–27]. We identified outcomes using clinical examination, pharmacy, medical procedure, and device codes to ensure accuracy, as these codes are often directly linked to insurance reimbursement [28,29]. If individuals were transferred to another facility after delivery, the outcomes were assessed using data from both the transferring and receiving facilities.

We defined PPH as follows: high-dose uterotonic use, blood transfusion, intra-uterine balloon tamponade, uterine rupture repair, trans-arterial embolization (TAE), and hysterectomy. High-dose uterotonic use was defined as either oxytocin ≥50 units or ≥3 types of uterotonics (oxytocin, methylergometrine, and prostaglandin E1 or F2α).

Infection was defined as ≥5 consecutive days of intravenous antibiotics use. A recent study defined serious infection as blood culture testing combined with ≥4 consecutive days of intravenous antibiotics [30]. We instead used ≥5 consecutive days of intravenous antibiotics because blood cultures are often unavailable at clinics, and nearly all patients receive antibiotic prophylaxis during CS [31].

Thrombosis was defined as a combination of clinical examinations (i.e., ultrasound or CT with contrast agents) and unfractionated heparin on the same day [32].

#### Area type

Healthcare provision is divided into 330 Medical Service Areas (MSAs) nationwide, of which 21 are located in Hokkaido. MSAs are classified as rural, provincial, or metropolitan based on population size and density (i.e., rural: population density <200 people/km^2^ and population <300,000; provincial: population density 200–1,000 people/km^2^ and population ≥300,000; metropolitan: population density ≥1,000 people/km^2^ and population ≥1,000,000) [33]. MSA classifications were obtained from publicly available data and visualized accordingly [19].

#### Maternal Comorbidity Index

The Maternal Comorbidity Index (MCI) was used to quantify maternal baseline comorbidity. The MCI is a weighted index that combines 20 conditions and maternal age [34]. We calculated the MCI from MHLW diagnosis codes, excluding “suspected flag”, during the covariate assessment window. To ensure positivity, we excluded comorbidities which were not managed at clinics (**Appendix Table 1**).

#### Other variables

Other variables included year of delivery and delivery mode (i.e., elective or emergent CS). Year of delivery was categorized as 2018–2019, 2020–2023, and 2024–2025 to account for the varying impact of the COVID-19 pandemic [35].

### Statistical analysis

Continuous variables were presented as mean (SD) or median (range) based on distribution, and categorical variables were presented as n (%).

We applied modified Poisson regression to estimate the risk ratios (RRs) for the association between facility type and the outcome [36]. Generalized estimating equations (GEEs) with an exchangeable working correlation structure were used to account for clustering within facilities [37]. For the two-category comparison, an interaction term between facility type and area type was included to account for regional variation. The three-category comparison, restricted to metropolitan areas, did not include this interaction term. The model was additionally adjusted for covariates, including the MCI (continuous), year of delivery (categorical), and delivery mode (elective or emergent CS).

We used g-computation to obtain standardized marginal probabilities of maternal complications, averaged over the distribution of other covariates within each area type. For each participant, predicted probabilities were derived from the fitted GEE model by counterfactually setting facility type while holding area type and all other covariates at their observed values. These predictions were then averaged within each area type to obtain standardized marginal probabilities for each facility type scenario. Confidence intervals for standardized marginal probabilities were obtained using the delta method, based on the robust variance-covariance matrix from the GEE model [38].

All analyses were conducted using R version 4.5.0 (R Foundation for Statistical Computing, Vienna, Austria). The statistical significance level was set at 0.05.

### Sensitivity analysis

We used an alternative definition of uterotonic use to examine the robustness of the threshold, as uterotonic use is a surrogate marker for PPH. High-dose uterotonic use was alternatively defined as oxytocin ≥60 units or ≥3 types of uterotonics.

## Results

Figure 2 shows the study flow. Among the 2,523 participants who underwent CS, 701 were excluded, resulting in 1,822 participants. Of these, 647 participants underwent CS in metropolitan areas. Area types in Hokkaido are visualized in Figure 3, showing 1 metropolitan area, 6 provincial areas, and 14 rural areas. Maternal demographics and outcomes are summarized in Table 1. A total of 1,027 participants underwent CS at PMCs, and 795 underwent CS at non-PMCs. The MCI scores were higher in the PMC group than in the non-PMC group. The incidence of maternal complications was 6.3% overall, 3.6% at PMCs, and 9.8% at non-PMCs. Although high-dose uterotonics were administered more frequently at non-PMCs, blood transfusion, intra-uterine balloon tamponade, and TAE were performed more frequently at PMCs. Infection was more frequently observed at non-PMCs. Thrombosis occurred in only one participant.

**Figure 2.**
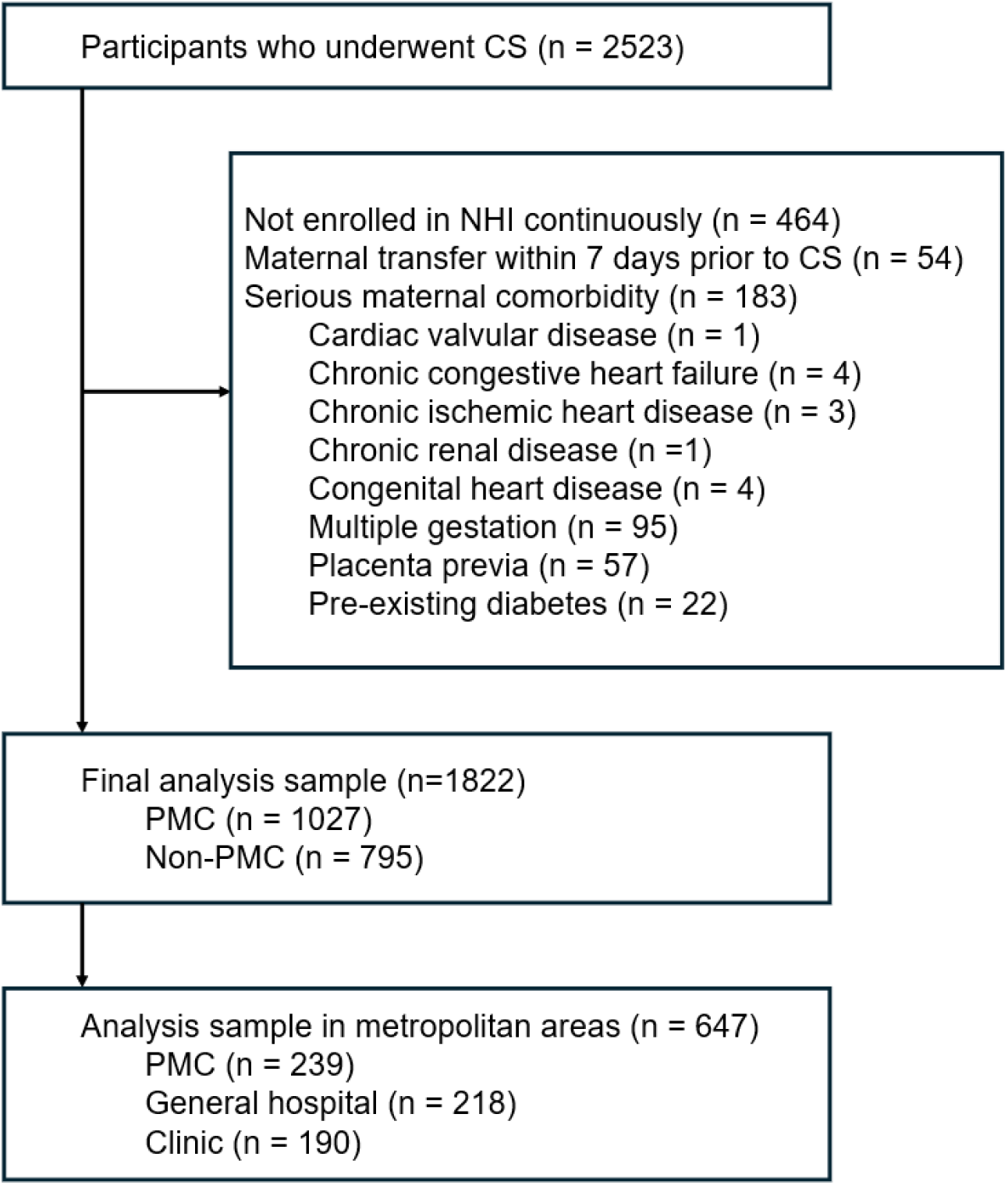
The study flow.

**Figure 3.**
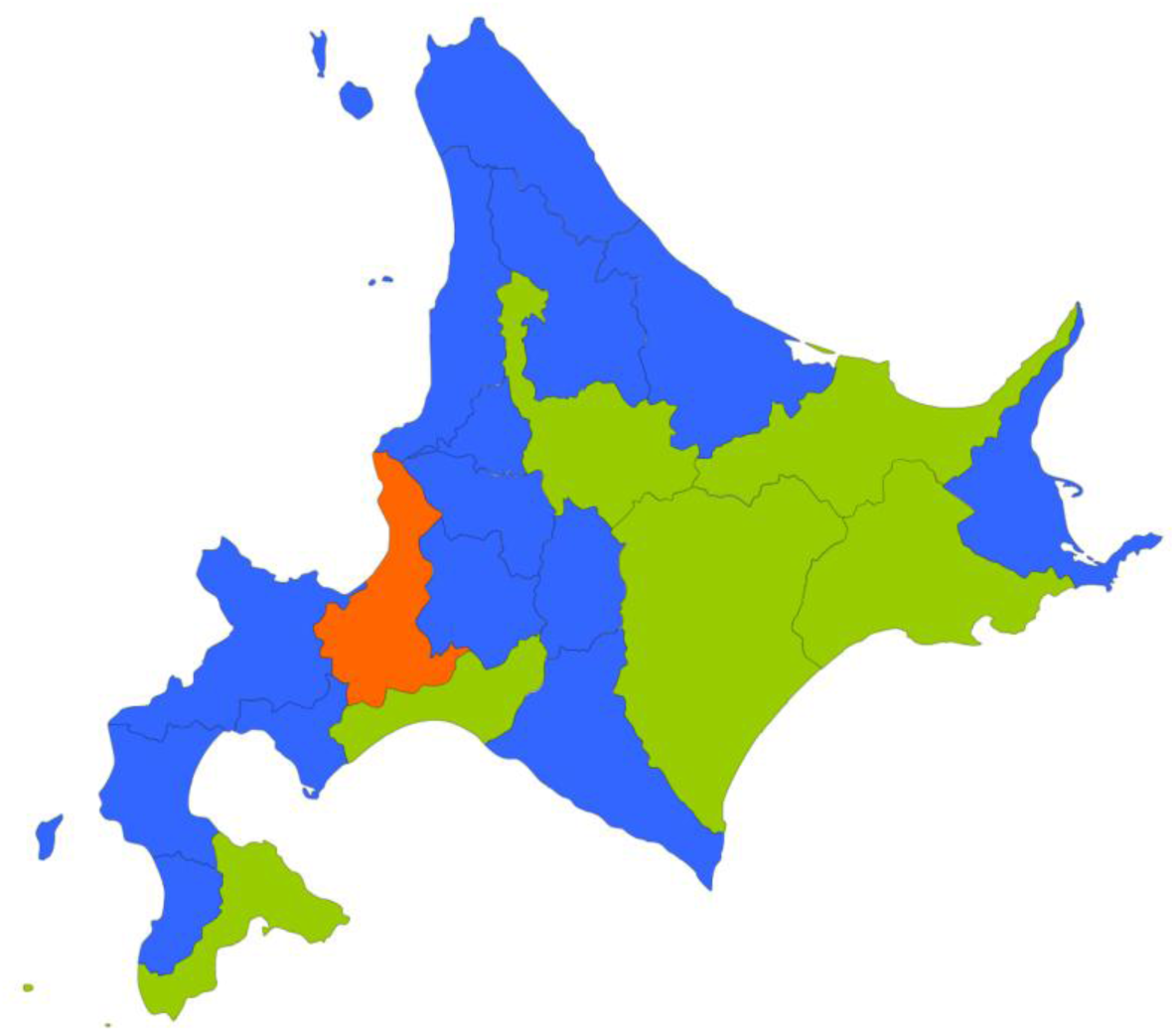
Area types in Hokkaido.

**Table 1.** Maternal demographics and outcomes.

Maternal demographics and outcomes
|  |  | **Overall**<br>N = 1822 | **PMC<sup>†</sup>**<br>N = 1027 | **Non-PMC<sup>†</sup>**<br>N = 795 | **p-value<sup>‡</sup>** |
| --- | --- | --- | --- | --- | --- |
| **Delivery mode** | n (%) |  |  |  | 0.86 |
| Elective cesarean |  | 1,089 (59.8%) | 612 (59.6%) | 477 (60.0%) |  |
| Emergent cesarean |  | 733 (40.2%) | 415 (40.4%) | 318 (40.0%) |  |
| **Region type** | n (%) |  |  |  | <0.001 |
| Rural area |  | 344 (18.9%) | 255 (24.8%) | 89 (11.2%) |  |
| Provincial area |  | 831 (45.6%) | 533 (51.9%) | 298 (37.5%) |  |
| Metropolitan area |  | 647 (35.5%) | 239 (23.3%) | 408 (51.3%) |  |
| **Year of delivery** | n (%) |  |  |  | 0.16 |
| 2018-2019 |  | 494 (27.1%) | 283 (27.6%) | 211 (26.5%) |  |
| 2020-2023 |  | 1,047 (57.5%) | 573 (55.8%) | 474 (59.6%) |  |
| 2024–2025 |  | 281 (15.4%) | 171 (16.7%) | 110 (13.8%) |  |
| **Maternal comorbidity index** | Median<br>[Min, Max] | 1 [0, 8] | 1 [0, 8] | 0 [0, 7] | <0.001 |
| **Maternal age** | n (%) |  |  |  |  |
| <35 |  | 1,000 (54.9%) | 522 (50.8%) | 478 (60.1%) |  |
| 35-39 |  | 600 (32.9%) | 365 (35.5%) | 235 (29.6%) |  |
| 40-44 |  | 214 (11.7%) | 136 (13.2%) | 78 (9.8%) |  |
| ≥45 |  | 8 (0.4%) | 4 (0.4%) | 4 (0.5%) |  |
| **Asthma** | n (%) | 28 (1.5%) | 16 (1.6%) | 12 (1.5%) |  |
| **Gestational hypertension** | n (%) | 19 (1.0%) | 15 (1.5%) | 4 (0.5%) |  |
| **Mild or unspecified preeclampsia** | n (%) | 64 (3.5%) | 50 (4.9%) | 14 (1.8%) |  |
| **Pre-existing hypertension** | n (%) | 25 (1.4%) | 17 (1.7%) | 8 (1.0%) |  |
| **Previous cesarean** | n (%) | 727 (39.9%) | 602 (58.6%) | 125 (15.7%) |  |
| **Severe preeclampsia** | n (%) | 62 (3.4%) | 56 (5.5%) | 6 (0.8%) |  |
| **Maternal complication** | n (%) | 115 (6.3%) | 37 (3.6%) | 78 (9.8%) |  |
| **Postpartum hemorrhage** | n (%) | 86 (4.7%) | 35 (3.4%) | 51 (6.4%) |  |
| Uterotonics | n (%) | 40 (2.2%) | 2 (0.2%) | 38 (4.8%) |  |
| Blood transfusion | n (%) | 40 (2.2%) | 26 (2.5%) | 14 (1.8%) |  |
| Intra-uterine balloon tamponade | n (%) | 11 (0.6%) | 10 (1.0%) | 1 (0.1%) |  |
| Trans-arterial embolism | n (%) | 5 (0.3%) | 3 (0.3%) | 2 (0.3%) |  |
| Uterine rupture repair | n (%) | 0 (0.0%) | 0 (0.0%) | 0 (0.0%) |  |
| Hysterectomy | n (%) | 1 (0.1%) | 0 (0.0%) | 1 (0.1%) |  |
| **Infection** | n (%) | 32 (1.8%) | 2 (0.2%) | 30 (3.8%) |  |
| **Thrombosis** | n (%) | 1 (0.1%) | 0 (0.0%) | 1 (0.1%) |  |

<sup>†</sup> PMC: Perinatal Medical Center, <sup>‡</sup> Categorical variables were compared using Pearson's Chi-squared test; continuous variables were compared using the Wilcoxon rank sum test.

The results from the GEEs are shown in Table 2. The risk of maternal complications was significantly lower at PMCs in rural and provincial areas (adjusted RR 0.37, 95% CI 0.16–0.88 in rural areas; adjusted RR 0.20, 95% CI 0.11–0.37 in provincial areas). In metropolitan areas as well, PMCs showed a lower risk for the outcome, though the association was not statistically significant (adjusted RR 0.75, 95% CI 0.39–1.44).

**Table 2.** Risk of maternal complications after cesarean section (the two-category comparison).

|  | PMC<sup>†</sup> | Non-PMC<sup>†</sup> | PMC<sup>†</sup> (vs. Non-PMC) |  |
| --- | --- | --- | --- | --- |
| Area type | Maternal complication risk (%) |  | Adjusted RR<sup>‡</sup> (95% CI) | p-value |
| **Rural** | 12/255 (4.7%) | 10/89 (11.2%) | 0.37 (0.16–0.88) | 0.024 |
| **Provincial** | 13/533 (2.4%) | 41/298 (13.8%) | 0.20 (0.11–0.37) | <0.001 |
| **Metropolitan** | 12/239 (5.0%) | 27/408 (6.6%) | 0.75 (0.39–1.44) | 0.384 |

<sup>†</sup> PMC: Perinatal Medical Center, <sup>‡</sup> RR: Risk Ratio; CI: Confidence Interval.

Standardized marginal probabilities of maternal complications are visualized in Figure 4, which shows higher risks at non-PMCs in rural and provincial areas.

**Figure 4.**
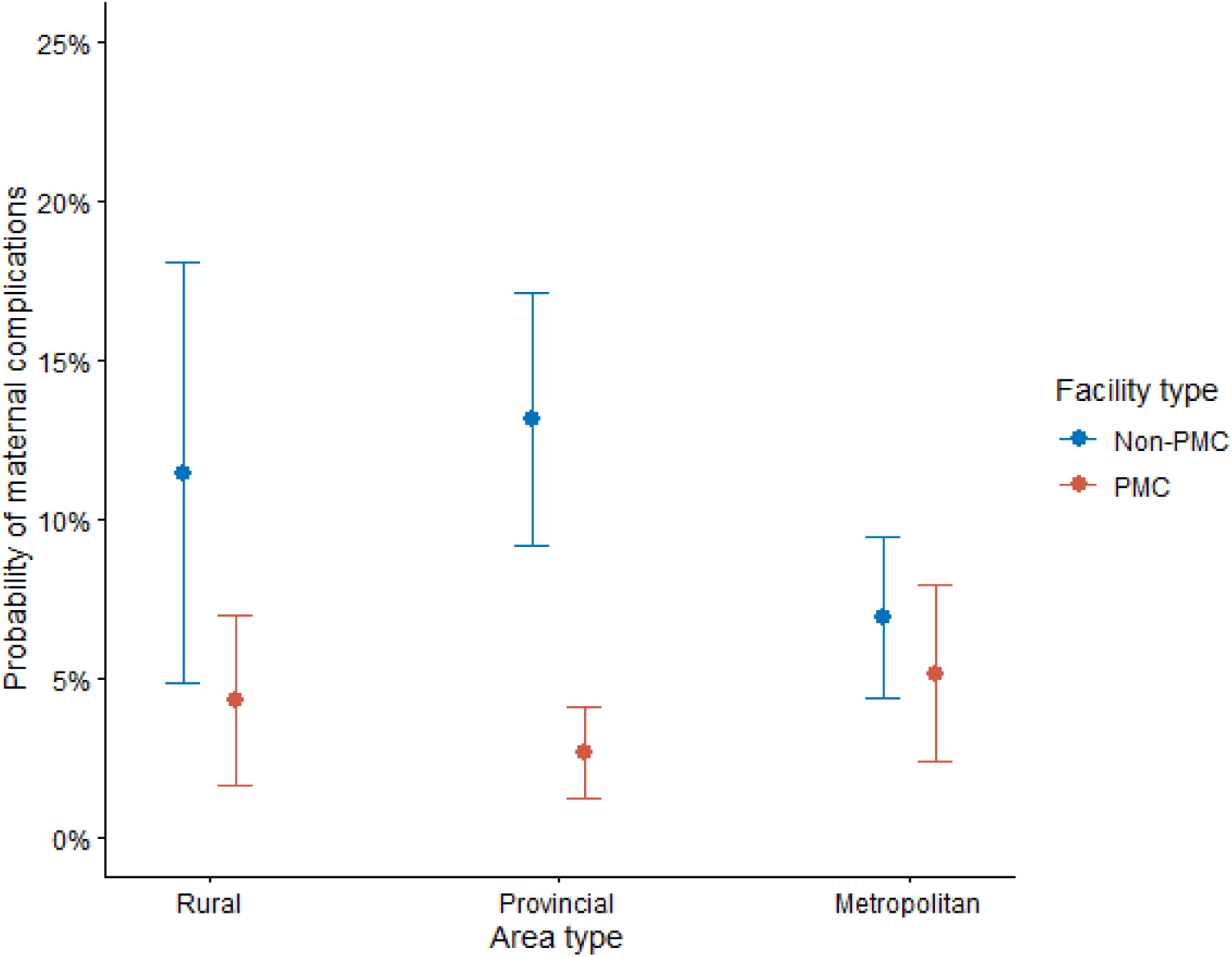
Standardized marginal probabilties of maternal complications across facility types.

Maternal demographics and outcomes in metropolitan areas, stratified by the three-category facility type, are summarized in **Appendix Table 7**. The three-category comparison is presented in Table 3, which favored PMCs and general hospitals over clinics (PMC vs clinic: adjusted RR 0.42, 95% CI 0.18–0.97; general hospital vs clinic: adjusted RR 0.25, 95% CI 0.09–0.70). Although PMCs had a higher risk than general hospitals, this association was not statistically significant (adjusted RR 1.66, 95% CI 0.56–4.91).

**Table 3.**
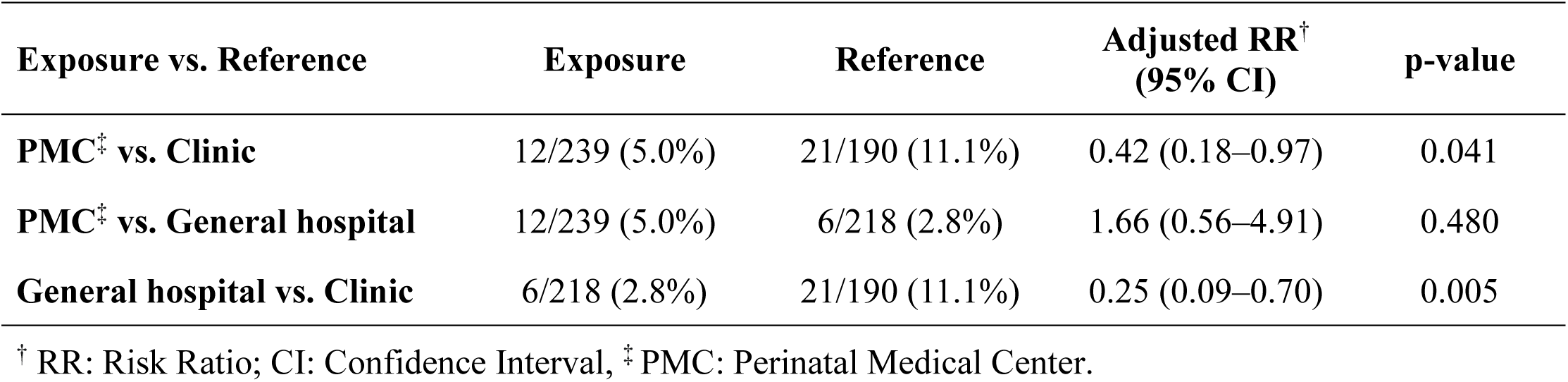
Risk of maternal complications after cesarean section in metropolitan areas (the three-category comparison).

Standardized marginal probabilities are presented in Figure 5, which shows higher risks at clinics.

**Figure 5.**
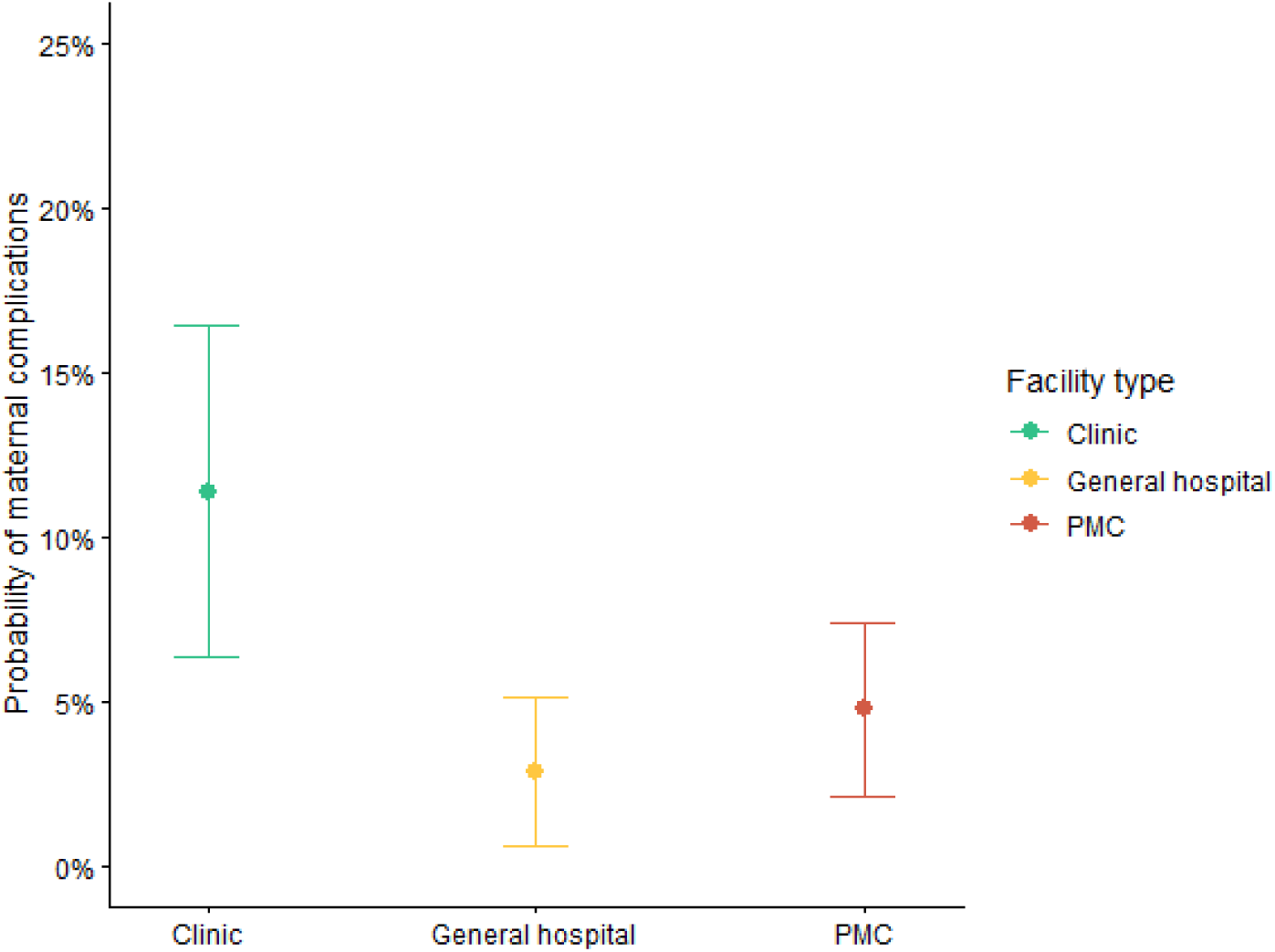
Standardized marginal probabilties of maternal complications across facility types in metropolitan areas.

### Sensitivity analysis

Results from the sensitivity analysis using an alternative threshold for the PPH definition are shown in **Appendix Tables 8–9**. The results were broadly consistent with the primary analysis, but the protective association of PMCs relative to non-PMCs in provincial areas was slightly attenuated compared with the primary analysis.

## Discussion

### Key results

In rural and provincial areas, CS performed at PMCs was associated with fewer maternal complications. In metropolitan areas, although the two-category comparison showed a lower risk ratio for PMCs, this association did not reach statistical significance. Nevertheless, the three-category comparison suggested that both PMCs and general hospitals had lower complication rates than clinics. Sensitivity analysis showed that the results were similar even when the threshold of uterotonics was changed, though the protective association in provincial areas was slightly attenuated compared with the primary analysis. Advanced interventions such as blood transfusion, intra-uterine balloon tamponade, and TAE were more frequently performed at PMCs.

### Interpretation

Our findings supported the advantage of higher-level facilities rather than higher-volume facilities, which was consistent with the previous study showing the advantage of higher hospital acuity [15]. The present study also highlighted regional variation in the association between facility type and maternal complications. While prior studies have reported regional variation in the association between other facility characteristics (e.g., volume or teaching status) and maternal complications [16,17], our findings extend this evidence by showing a similar regional pattern with respect to facility type.

The present study suggests risks associated with CS at low-level facilities in Japan. Recent studies reported that while blood transfusion was more frequently performed at higher-level facilities (i.e., PMCs and higher-level general hospitals) [39], approximately 90% of maternal deaths due to PPH occurred at lower-level facilities [40]. Additionally, over half of maternal mortality was accompanied by maternal transfer from lower-level facilities to higher-level facilities [21]. These studies suggest that lower-level facilities may predispose patients to serious maternal consequences, such as mortality, even under a risk-based role differentiation system. The present study contributes a direct comparison of maternal complications after CS among these facility types.

The higher risks observed after CS at non-PMCs in rural and provincial areas, and at clinics in metropolitan areas, may reflect staffing constraints. Obstetricians are maldistributed in Japan, with fewer practicing at non-PMCs in rural and provincial areas [12]. A similar staffing constraint applies to clinics in metropolitan areas, most of which are run by ≤2 obstetricians [10,21]. This staffing pattern highlights the vulnerability of these facilities in managing obstetric emergencies [21,39].

Regional variations may also reflect a constrained referral network. In metropolitan areas, PMCs, general hospitals, and clinics are located close to one another, whereas in rural and provincial areas they are often far apart [11,33]. This geographical barrier constrains risk-based role differentiation in rural and provincial areas, where non-PMCs sometimes manage high-risk pregnancies because referral is not a realistic option.

The higher risks observed at PMCs relative to general hospitals in metropolitan areas may reflect residual confounding rather than differences in quality of care. Similar findings were reported at extremely high-volume facilities, where higher maternal complication rates were attributed to high patient volume, insufficient supervision of trainees, and unmeasured case severity [7,8]. Likewise, the NHI data lack several potential confounders, including medical comorbidities (e.g., maternal body mass index, number of previous cesarean sections, gestational weeks at delivery), socioeconomic status, and health literacy. The unmeasured confounding may have inflated the standardized marginal probabilities of maternal complications at PMCs.

Our results are particularly relevant with respect to regional consolidation of obstetric care. In rural and provincial areas, consolidating deliveries into PMCs may be warranted from a patient safety perspective. In metropolitan areas, where PMCs, general hospitals, and clinics are frequently all within reach, the higher risks at clinics warrant greater public attention, as these risks are not widely recognized by patients. Given the broader choice of facility type available to patients in metropolitan areas, informing them of the risks at clinics could encourage safer decisions regarding delivery facilities.

### Strengths and limitations

The strengths of the present study include the use of the NHI claims data across diverse areas in Hokkaido, which enabled comparisons across facility types while also accounting for regional variations. Additionally, these data allowed patient tracking even when individuals were transferred to another facility. Sensitivity analysis also showed the robustness of our findings to alternative definitions of uterotonic use. Although the association between facility type and complications was somewhat attenuated at non-PMCs in provincial areas, the overall conclusions remained unchanged. Finally, we excluded participants with serious comorbidities, which contributed to comparability of populations across facility levels.

However, several limitations should be acknowledged. First, the present study has limited generalizability to populations with different insurance status or to other regions. In addition, we only investigated CS because it is covered by insurance, whereas normal delivery (i.e., vaginal delivery requiring no medical intervention) is not covered and thus does not appear in the NHI data. Moreover, our findings reflect how maternal complications are managed rather than their clinical diagnosis. In particular, uterotonic use served as a surrogate marker for PPH and thus may reflect differences in clinical protocols rather than true differences in PPH incidence. The use of the MCI also raises concerns regarding its validity. Although the MCI has been validated using Asian populations [41], it has not been validated using Japanese insurance claims data, which could potentially lead to misclassification.

### Conclusions

Maternal complications following CS occurred less frequently at PMCs than at non-PMCs in rural and provincial areas. In metropolitan areas, CS at PMCs and general hospitals was associated with lower risks compared to clinics. These findings should be considered for regional consolidation of obstetric care.

## Data Availability

All data produced in the present study are available upon reasonable request to the authors.

## Acknowledgement

This research is supported by KAKENHI (grant No. 22K15662 to KA) from the Japan Society for the Promotion of Science. TY is supported by Graduate Scholarship for Degree Seeking Students by the Japan Student Services Organization (JASSO) Student Exchange Support Program and Global Grant Scholarship by the Rotary Foundation (grant No. GG2579376).

## Figure legends

Figure 1. Graphical depiction of study design. NHI: national health insurance.

Figure 2. Inclusion and exclusion of study participants. CS: cesarean section; NHI: national health insurance; PMC: perinatal medical center.

Figure 3. Area types are visualized in Hokkaido. Orange: metropolitan areas; Green: provincial areas; Blue: rural areas.

Figure 4. Standardized marginal probabilities of maternal complications are shown for each facility type and area type. PMC: perinatal medical center.

Figure 5. Standardized marginal probabilities of maternal complications are shown for each facility level in metropolitan areas. PMC: perinatal medical center.

