## Appendix for "Maternal complications after cesarean section by obstetric facility type in Japan: A claims-based cohort study"

**Appendix Table 1.**
**The maternal comorbidity index**

| Comorbidity | Weight |
|---|---|
| Alcohol abuse† | 1 |
| Asthma | 1 |
| Cardiac valvular disease† | 2 |
| Chronic congestive heart failure† | 5 |
| Chronic ischemic heart disease† | 3 |
| Chronic renal disease† | 1 |
| Congenital heart disease† | 4 |
| Drug abuse† | 2 |
| Gestational hypertension | 1 |
| HIV† | 2 |
| Maternal age | |
| 35-39 | 1 |
| 40-44 | 2 |
| $\geq 45$ | 3 |
| Mild or unspecified preeclampsia | 2 |
| Multiple gestation† | 2 |
| Placenta previa† | 2 |
| Pre-existing diabetes† | 1 |
| Pre-existing hypertension | 1 |
| Previous cesarean | 1 |
| Pulmonary hypertension† | 4 |
| Severe preeclampsia | 5 |
| Sickle cell disease† | 3 |
| Systemic lupus erythematosus† | 2 |

† These conditions were excluded because they were not observed at clinics. A participant's comorbidity index is generated by summing the weights of all maternal comorbidities during the covariate assessment window.

**Appendix Table 2.**
**MHLW† codes for clinical examinations**

| Examination | MHLW† codes |
|---|---|
| CT | 170011710, 170011810, 170012110, 170028610, 170033410, 170034910, 170038710, 170038810, 170038910, 170039010, 170039110, 170040210, 170040310, 170040410, 170040510, 170040610, 170040710, 170040810, 170041010, 170041110 |
| Ultrasonography for thrombosis | 160213010 |

† MHLW: Ministry of Health, Labour and Welfare

**Appendix Table 3.**
**MHLW† codes for pharmacy**

| **Pharmacy** | **MHLW† codes** |
|---|---|
| Contrast agent for CT | 621782301, 620004304, 621184501, 621451401, 621185901, 620009145, 621189001, 620003695, 621185301, 620003692, 620007447, 621187501, 621183701, 620003689, 620003684, 621184101, 621191201, 620009149, 621929103, 621453402, 621928503, 620003686, 621183301, 621892301, 621188701, 620009147, 621186501, 621728902, 621452302, 621188901, 620003694, 620003685, 621190101, 621534901, 620007448, 620007449, 621176405, 621188601, 621451501, 620003691, 621184602, 621190901, 621190501, 647210005, 621191901, 621188001, 621188801, 621928803, 620003697, 620009146, 621191701, 620003688, 621453002, 621191101, 620009142, 621929003, 620003690, 620007451, 621453202, 621185802, 622547001, 620009148, 620009144, 622213602, 621190801, 621695401, 621185703, 622766000, 621184901, 621189901, 621183802, 621758502, 621186103, 621452202, 621922401, 621452002, 620004305, 621190701, 621183101, 621452702, 621191001, 621928203, 621928403, 621187901, 622028102, 621190201, 622376902, 622067803, 620003682, 622067901, 621184202, 622766400, 621453302, 621490701, 622765800, 622179401, 622030802, 621453102, 620007450, 621190301 |
| Fresh frozen plasma | 621772701, 622192101, 621772601 |
| Intravenous antibiotics | 620002955, 620002977, 620003002, 620003003, 620003004, 620003210, 620003733, 620003734, 620003735, 620003736, 620003737, 620003738, 620003739, 620003740, 620003815, 620003816, 620003817, 620004134, 620004135, 620004151, 620004155, 620004652, 620004653, 620004775, 620004776, 620005201, 620005202, 620005674, 620005675, 620005676, 620006243, 620006244, 620006316, 620006317, 620007318, 620007365, 620007514, 620007518, 620007519, 620007520, 620008446, 620008447, 620009563, 620009586, 620009588, 621076601, 621076801, 621077201, 621077501, 621078106, 621078301, 621078601, 621078906, 621079101, 621079401, 621096002, 621096601, 621096702, 621096802, 621097401, 621097602, 621102102, 621102802, 621111802, 621111803, 621112003, 621112004, 621114602, 621115302, 621344901, 621345301, 621347901, 621441901, 621488403, 621488504, 621488601, 621538203, 621538303, 621540002, 621555204, 621703001, 621703101, 621703201, 621708501, 621727601, 621756202, 621756304, 621756603, 621757001, 621766301, 621766401, 621812201, 621931801, 621946701, 621946801, 621947901, 621948001, 621951701, 621952301, 621952401, 621952501, 621966801, 621966901, 621967202, 621967801, 621968001, 621987301, 621987401, |

| | |
|---|---|
| | 621987501, 621987602, 621994601, 621994701, 622033402, 622037301, 622045702, 622051702, 622052901, 622074802, 622077303, 622078501, 622079701, 622079801, 622079901, 622080002, 622099301, 622100601, 622111401, 622118402, 622124801, 622124902, 622125001, 622130801, 622131001, 622131101, 622419401, 622419601, 622423101, 622439001, 622441301, 622450601, 622450701, 622453701, 622453801, 622453901, 622454001, 622458201, 622460701, 622460801, 622460901, 622467201, 622470601, 622470701, 622470801, 622470901, 622471301, 622669501, 622763600, 622763700, 622764100, 622764200, 622764500, 622764600, 622764700, 622764800, 622949701, 622949801, 622949901, 622950001, 622961200, 622961300, 622961400, 622970901, 622983700, 622983800, 640407080, 640407081, 640408148, 640408149, 640443048, 640444050, 640444071, 640451036, 640451037, 640454018, 640454019, 640462059, 640462060, 640470011, 646120011, 646120012, 646130037, 646130072, 646130073, 646130074, 646130075, 646130076, 646130121, 646130122, 646130123, 646130124, 646130136, 646130137, 646130264, 646130265, 646130268, 646130269, 646130301, 646130302, 646130368, 646130369, 670008446, 670008447 |
| Methylergometrine | 620006280, 620559802, 642520008 |
| Oxytocin | 620003123, 620006611, 642410006 |
| Prostaglandin E1 or F2α | 620553801, 642490022 |
| Red blood cell | 621772101, 621772901, 622191601, 622191801, 620004680, 621772001, 621772801 |
| Unfractionated heparin | 620006725, 620006728, 620006734, 620006739, 620812504, 620812701, 621825302, 621825502, 621825602, 621825704, 621825802, 621826004, 621826102, 621826402, 622757800, 622757900, 622973301, 643330011 |

† MHLW: Ministry of Health, Labour and Welfare

**Appendix Table 4.**
**MHLW† codes for medical procedures**

| Procedure | MHLW† codes |
|---|---|
| Elective cesarean section | 150222210 |
| Emergent cesarean section | 150222110 |
| Hysterectomy | 150217410, 150217510, 150223110, 150222810 |
| Maternal transfer | 190126910 |
| Trans-arterial embolism | 150360610, 150360710, 150376810 |
| Uterine rupture repair | 150223010 |

† MHLW: Ministry of Health, Labour and Welfare

**Appendix Table 5.**
**MHLW† codes for devices**

| Device | MHLW† codes |
| --- | --- |
| Uterine balloon tamponade | 710010719 |

† MHLW: Ministry of Health, Labour and Welfare

**Appendix Table 6.**
**MHLW† codes for diagnosis**

| Diagnosis | MHLW† codes |
|---|---|
| Alcohol abuse | 3050006, 2914002, 3039002, 3050001, 2914001, 8830339, 8830330, 8830355, 8830331, 8830345, 8849011, 2940005, 8842547, 8830357, 2919003, 8849010, 8830352, 3050004, 2919005, 8830798, 2911002, 8830344, 8840299, 8830337, 8845631, 8830349, 8830346 |
| Asthma | 4912003, 4912004, 4912001, 8840399, 8850253, 8849096, 8849097, 8849192, 8849212, 8849256, 8846176, 4939008, 4930005, 8847408, 8830247, 4939037, 4939038, 4930001, 4930002, 8833884, 4930006, 4939016, 4939039, 4939022, 4939003, 8841641, 8831609, 8844994, 8834797 |
| Cardiac valvular disease | 4240018, 3949001, 8836698, 4240009, 8836695, 8836697, 3949003, 8840945, 8840946, 3942006, 8840949, 8849007, 8840950, 8840948, 3970005, 3970011, 3970008, 8834110, 8834111, 8834113, 8840940, 8840939, 8840941, 8834109, 4249020, 8848942, 8848943, 8848945, 8848946, 8848958, 8848960, 8848879, 8848880, 8848944, 8848947, 8848959, 8848962, 8848965, 8848967, 8848881, 8848948, 8848966, 8848969, 8848964, 8848971, 8848972, 8848940, 8848956, 8848941, 8848957, 8848963, 8848968, 8848970, 8848961, 8840942, 8840943, 3980001, 3989002, 3989003, 8840944, 8840411, 3979001, 8840954, 8840410, 8840412, 8840409, 7469018, 3949004, 8842518, 3949002, 4240019, 8836699, 4240004, 8839422, 8849668, 8845179, 4241009, 4241015, 8837399, 4241025, 8837402, 8837403, 8839423, 8843935, 8839420, 8839419, 8839421, 8839418, 4243015, 8838864, 8838867, 8838863, 8840043, 8835117, 4249004, 8840044, 8833133 |
| Chronic congestive heart failure | 4289015, 8842461, 4280011, 8834012, 4281009, 8834931, 4281005, 8841016, 4280002, 4289018, 4289005, 8830796, 5140016, 8851083, 4281010, 4280005, 8851575, 8851085, 8851086, 8851084 |
| Chronic ischemic heart disease | 4139007, 4139004, 4139028, 8847838, 8830403, 4139026, 8841212, 8847005, 8845119, 8834877, 8836816, 8844586, 8841211, 4141004, 8842762, 4148001, 4140014, 8831573, 4141003, 8846370, 8845239, 4149007, 8842760, 4141001, 8832678, 8837807, 4140020, 4140008, 8842211, 4110003, 8831574, 8831577, 4140013, 8837801, 8849595, 8837804, 8847530, 8837810, 8831572, 8837809 |
| Chronic renal disease | 5829003, 5829008, 8840376, 8832919, 8840393, 8840391, 8840392, 8840394, 5819004, 8834799, 8838401, 8849711, 8838367, 8849828, 8836335, 8839471, 8848070, 8839430, 8835738, 8837974, 8849833, 8839551, 8850255, 8850591, 8839535, 5839010, 5831004, 5839007, 8848410, 8836667, 8836820, 8840229, 8838994, 8831660, 8850032, 8840538, 8840230, 8840231, 8840232, 8849712, 8835611, 8835636, 8842116, 5859002, |

| | |
|---|---|
| | 5869016, 3621014, 8844106, 8841385, 5869015, 8847544, 8838555, 8847577, 8847578, 8847579, 8847582, 8847583, 8847580, 8847581, 8848103, 8851366, 8838554, 8851031, 8847501, 8835613, 8847972, 5881001, 8835615, 5880002, 2762015, 8832417, 8830899, 8832721, 8848582, 8838517, 8838518, 8840870, 8833311, 8838396, 8841309, 8830501, 8847502, 8844495, 5870001, 4039015, 4039018 |
| Congenital heart disease | 8834017, 8835002, 7450002, 8837116, 8834985, 8837268, 8830784, 8834011, 7451011, 8848826, 7460012, 8848253, 8848599, 8831636, 8842798, 7451001, 7451013, 8848480, 8848481, 8848482, 7454005, 8830500, 8837117, 7456003, 8839591, 8839593, 8851544, 8839592, 8845263, 8848715, 8840885, 8835111, 8834988, 8835119, 8835132, 8837396, 7456007, 7454010, 8848790, 8831634, 8841660, 8850075, 7454008, 8839631, 8841661, 7454009, 8850017, 8844147, 8848553, 8848691, 8848464, 8834112, 8838866, 8845467, 7469022, 8830793, 8838865, 8848729, 8836222, 8844500, 8836346, 8836347, 7472013, 7464007, 8838403, 7466004, 8849648, 8834015, 8836279, 8836278, 8836280, 8836295, 8836296, 8836277, 7468002, 8830792, 7468001, 8833993, 8834014, 7469016, 7468007, 7468046, 8835143, 8847160, 8849390, 7469020, 7468029, 8835134, 8848746, 8830576, 7469008, 8842676, 7469006, 8836235, 8848605, 8849261, 8836220, 8836232, 8836236, 8850667, 8836155, 8836237, 8850665, 8838862, 7468005, 8836154, 4251001, 8837459, 7468045, 8849467, 8849442, 8849443, 8849468, 8849743, 8850430, 7472024, 7473005, 7473019, 8842554, 8842677, 8838855, 7473008, 8849685, 8837392, 8838259, 8849674, 8835310, 8848269, 8837389, 8837394, 7472023, 8837401, 8838856, 7472001, 7478021, 8836348, 8837388, 8849661, 8837264, 8836345, 8849808, 8849660, 8837390, 8838857, 8836294, 8836344, 8838861, 8849684, 8849554, 8842803, 8843759, 8849673, 7471011, 8843791, 8849444, 8849686, 8842742, 8849675, 8848554, 7474026, 4529005, 7476023, 7476008, 7474027, 8842259, 7476036, 7511028, 7476034, 7476033, 8838839, 7476035, 8840690, 8837263, 8849804, 8836573, 8842790, 8838836, 8842848, 7474014, 8831370, 8836291, 8849377, 8836150, 8836245, 7474025, 7474022, 7474024, 7474023 |
| Drug abuse | 3055001, 3040001, 3040004, 3055003, 3040003, 3055002, 8844834, 8840285, 3043002, 8849155, 8844895, 8849156, 3041003, 3041001, 3041004, 3046006, 8843968, 8844901, 3042001, 3056001, 8844870, 8831428, 8830408, 8831430, 3057001, 8830410, 8844839, 3046007, 8844840, 8846493, 8844843, 8844835, 8844844, 3053001, 3045004, 3053003, 8844833, 8844864, 3046004, 3046005, 3046001, 3046003, 3046008, 3046002, 8844850, 3049002, 3059002, 3059003, 3058002, 2940003, 2922001, 2949013, 2920003, 2929001, 2949012, 3049004, 8844912, 8844896, 8844879 |

| | |
|---|---|
| Gestational hypertension | 6429003, 8838596, 8834071 |
| HIV | 8830092, 8830094, 8850707, 8849060, 8843639, 8830096, 8850708, 2793011, 8842156, 0798002, 8847287, 8847288, 7712015, 8830055, 2793007, 8830095 |
| Mild or unspecified preeclampsia | 8842687, 8842828, 8842827, 8848335, 8842709, 8842804, 8842791 |
| Multiple gestation | 6518001, 6512001, 6519001, 6511001, 6510003, 8845591, 8845518, 8845517, 8836991, 8851445, 8851438, 8851446, 8851439, 8851444, 8851437, 8851447, 8851440 |
| Placenta previa | 6411001, 6410002, 6411007, 6411005, 6567002, 6411003, 6411006, 6411004, 8834646, 8834647 |
| Pre-existing diabetes | 2500014, 2500027, 8843105, 8844045, 8845046, 8830030, 8844626, 8845043, 8849056, 8845054, 8830031, 8845059, 8844022, 8845044, 8845049, 8845053, 8845056, 8845842, 8843982, 8845047, 8845048, 8845061, 8830032, 8844346, 8845068, 8830033, 8830028, 8845058, 8845045, 8845050, 8845051, 8843983, 8843984, 8843985, 8843988, 8843989, 8845060, 8845063, 8841679, 8843986, 8843987, 8845062, 8841682, 8841681, 8845066, 8844627, 8845055, 8845065, 8845067, 8845069, 8841685, 8841680, 8845052, 8845057, 8845064, 8845070, 8845071, 8841683, 8844026, 8844024, 8844028, 8841686, 8841688, 8841684, 8851450, 8844023, 8851474, 8844025, 8851432, 8849557, 8841687, 8844536, 8844027, 8844030, 8844031, 8844029, 2500015, 8830405, 8843106, 8835244, 8845075, 8830041, 8844628, 8845072, 8849058, 8845083, 8830042, 8845088, 8830044, 2500001, 8845073, 8845078, 8845082, 8845085, 8848108, 8843990, 8845076, 8845077, 8845090, 8830043, 8844347, 8845097, 8830045, 8830040, 8845087, 8845074, 8845079, 8845080, 8843991, 8843992, 8843993, 8843996, 8843997, 8845089, 8845092, 8841689, 8843994, 8843995, 8845091, 8841692, 8841691, 8845095, 8844629, 8845084, 8845094, 8845096, 8845098, 8841695, 8841690, 8845081, 8845086, 8845093, 8845099, 8845100, 8841693, 8841696, 8841698, 8841694, 8851451, 8851475, 8851433, 8849558, 8841697, 8844537, 8830029, 8830039, 8838621 |
| Pre-existing hypertension | 8833421, 8849300, 8830212, 8842488, 4019016, 8840107, 8832479, 8842178, 8842089, 8842500, 8842094, 8848337, 4019017, 8851401, 8833426, 4029010, 8851400, 8833422, 4039025, 4039001, 4039006, 8833427, 5879003, 4039005, 4039033, 4039026, 4039036, 4039028, 8833425, 8849491, 8849279, 8835614, 8838398, 8839689, 8838336, 8835586, 8835605, 8845461, 8845458, 8845460, 8845462, 8845459 |

| | |
|---|---|
| Previous cesarean section | 6542002, 8832140, 8832141, 8851463, 6542003, 8839049, 8844756 |
| Pulmonary hypertension | 4169002, 4169003, 4169004, 8838825, 8830160, 8844798, 8845466, 8846206, 8844804, 8841669, 8846195 |
| Severe preeclampsia | 6426003, 6426006, 6426007, 6426008, 6426001, 8851621, 8842091, 8845716, 6681001, 8845665, 8848093, 8848100, 8848054, 8842155, 8848357, 8842765 |
| Sickle cell disease | 8834025, 8832782, 8840024, 8830369, 8834026, 8837559, 8837965, 8830572, 8839966, 8839967, 8839968, 8839969, 8831451, 8831453, 8839956, 8831452, 8835314, 8836819 |
| Systemic lupus erythematosus | 8830137, 7100007, 8844341, 8844340, 8850360, 8844342, 8844339, 8844080, 7100011, 7100031, 8840979, 8836519, 8836518, 8842174, 8841438, 8836515, 8836520, 8846167, 8836516, 8848278, 8836513 |

† MHLW: Ministry of Health, Labour and Welfare

**Appendix Table 7.**
**Maternal demographics and outcomes in metropolitan areas**

| | | **Overall** N = 647 | **PMC**† N = 239 | **General hospital** N = 218 | **Clinic** N = 190 | **p-value**‡ |
|---|---|---|---|---|---|---|
| **Delivery mode** | n (%) | | | | | 0.18 |
| Elective cesarean | | 384 (59.4%) | 132 (55.2%) | 139 (63.8%) | 113 (59.5%) | |
| Emergent cesarean | | 263 (40.6%) | 107 (44.8%) | 79 (36.2%) | 77 (40.5%) | |
| **Year of delivery** | n (%) | | | | | 0.18 |
| 2018-2019 | | 174 (26.9%) | 66 (27.6%) | 51 (23.4%) | 57 (30.0%) | |
| 2020-2023 | | 389 (60.1%) | 137 (57.3%) | 145 (66.5%) | 107 (56.3%) | |
| 2024–2025 | | 84 (13.0%) | 36 (15.1%) | 22 (10.1%) | 26 (13.7%) | |
| **Maternal comorbidity index** | Median [Min, Max] | 1 [0, 7] | 2 [0, 7] | 1 [0, 7] | 0 [0, 6] | <0.001 |
| **Maternal age** | n (%) | | | | | |
| <35 | | 354 (54.7%) | 97 (40.6%) | 135 (61.9%) | 122 (64.2%) | |
| 35-39 | | 212 (32.8%) | 99 (41.4%) | 58 (26.6%) | 55 (28.9%) | |
| 40-44 | | 76 (11.7%) | 41 (17.2%) | 23 (10.6%) | 12 (6.3%) | |
| ≥45 | | 5 (0.8%) | 2 (0.8%) | 2 (0.9%) | 1 (0.5%) | |
| **Asthma** | n (%) | 8 (1.2%) | 5 (2.1%) | 3 (1.4%) | 0 (0.0%) | |

| | | Overall N = 647 | PMC† N = 239 | General hospital N = 218 | Clinic N = 190 | p-value‡ |
|---|---|---|---|---|---|---|
| **Gestational hypertension** | n (%) | 2 (0.3%) | 1 (0.4%) | 1 (0.5%) | 0 (0.0%) | |
| **Mild or unspecified preeclampsia** | n (%) | 20 (3.1%) | 13 (5.4%) | 6 (2.8%) | 1 (0.5%) | |
| **Pre-existing hypertension** | n (%) | 6 (0.9%) | 3 (1.3%) | 2 (0.9%) | 1 (0.5%) | |
| **Previous cesarean** | n (%) | 199 (30.8%) | 134 (56.1%) | 48 (22.0%) | 17 (8.9%) | |
| **Severe preeclampsia** | n (%) | 20 (3.1%) | 15 (6.3%) | 4 (1.8%) | 1 (0.5%) | |
| **Maternal complication** | n (%) | 39 (6.0%) | 12 (5.0%) | 6 (2.8%) | 21 (11.1%) | |
| **Postpartum hemorrhage** | n (%) | 28 (4.3%) | 12 (5.0%) | 6 (2.8%) | 10 (5.3%) | |
| Uterotonics | n (%) | 9 (1.4%) | 0 (0.0%) | 4 (1.8%) | 5 (2.6%) | |
| Blood transfusion | n (%) | 16 (2.5%) | 9 (3.8%) | 2 (0.9%) | 5 (2.6%) | |
| Intra-uterine balloon tamponade | n (%) | 6 (0.9%) | 5 (2.1%) | 0 (0.0%) | 1 (0.5%) | |
| Trans-arterial embolism | n (%) | 3 (0.5%) | 2 (0.8%) | 0 (0.0%) | 1 (0.5%) | |
| Uterine rupture repair | n (%) | 0 (0.0%) | 0 (0.0%) | 0 (0.0%) | 0 (0.0%) | |
| Hysterectomy | n (%) | 0 (0.0%) | 0 (0.0%) | 0 (0.0%) | 0 (0.0%) | |

| | | Overall<br>N = 647 | PMC[†]<br>N = 239 | General hospital<br>N = 218 | Clinic<br>N = 190 | p-value[‡] |
|---|---|---|---|---|---|---|
| **Infection** | n (%) | 11 (1.7%) | 0 (0.0%) | 0 (0.0%) | 11 (5.8%) | |
| **Thrombosis** | n (%) | 0 (0.0%) | 0 (0.0%) | 0 (0.0%) | 0 (0.0%) | |

[†] PMC: Perinatal Medical Center, [‡] Categorical variables were compared using Pearson's Chi-squared test; continuous variables were compared using the Wilcoxon rank sum test.

**Appendix Table 8.**

**Risk of maternal complications after cesarean section using the alternative threshold (the two-category comparison)**

| | PMC† | Non-PMC† | PMC† (vs. Non-PMC) | |
|---|---|---|---|---|
| **Area type** | **Maternal complication risk (%)** | | **Adjusted RR‡ (95% CI)** | **p-value** |
| **Rural** | 12/255 (4.7%) | 10/89 (11.2%) | 0.38 (0.16–0.88) | 0.025 |
| **Provincial** | 13/533 (2.4%) | 15/298 (5.0%) | 0.51 (0.24–1.06) | 0.071 |
| **Metropolitan** | 12/239 (5.0%) | 25/408 (6.1%) | 0.80 (0.41–1.56) | 0.504 |

† PMC: Perinatal Medical Center, ‡ RR: Risk Ratio; CI: Confidence Interval.

**Appendix Table 9.**

**Risk of maternal complications after cesarean section in metropolitan areas using the alternative threshold (the three-category comparison)**

| Exposure vs. Reference | Exposure | Reference | Adjusted RR[†] (95% CI) | p-value |
|---|---|---|---|---|
| **PMC[‡] vs. Clinic** | 12/239 (5.0%) | 20/190 (10.5%) | 0.42 (0.18–1.00) | 0.051 |
| **PMC[‡] vs. General hospital** | 12/239 (5.0%) | 5/218 (2.3%) | 1.94 (0.60–6.21) | 0.350 |
| **General hospital vs. Clinic** | 5/218 (2.3%) | 20/190 (10.5%) | 0.22 (0.07–0.66) | 0.004 |

[†] RR: Risk Ratio; CI: Confidence Interval, [‡] PMC: Perinatal Medical Center.
